# Can the protection be among us? Previous viral contacts and prevalent HLA alleles avoiding an even more disseminated COVID-19 pandemic

**DOI:** 10.1101/2020.06.15.20131987

**Authors:** Eduardo Cheuiche Antonio, Mariana Rost Meireles, Marcelo Alves de Souza Bragatte, Gustavo Fioravanti Vieira

## Abstract

COVID-19 is bringing scenes of sci-fi movies into real life, and it seems to be far from over. Infected individuals exhibit variable severity, suggesting the involvement of the genetic constitution of populations and previous cross-reactive immune contacts in the individuals’ disease outcome. To investigate the participation of MHC alleles in COVID-19 severity, the combined use of HLA-B*07, HLA-B*44, HLA-DRB1*03, and HLA-DRB1*04 grouped affected countries presenting similar death rates, based only on their allele frequencies. To prospect T cell targets in SARS-CoV-2, we modeled 3D structures of HLA-A*02:01 complexed with immunogenic epitopes from SAR-CoV-1 and compared them with models containing the corresponding SARS-CoV-2 peptides. It reveals molecular conservation between SARS-CoV peptides, evidencing that the corresponding current sequences are putative T cell epitopes. These structures were also compared with other HCoVs sequences, and with a panel of epitopes from unrelated viruses, looking for the triggers of cross-protection in asymptomatic and uninfected individuals. 229E, OC43, and impressively, viruses involved in endemic human infections share fingerprints of immunogenicity with SARS-CoV peptides. Wide-scale HLA genotyping in COVID-19 patients shall improve prognosis prediction. Structural identification of previous triggers paves the way for herd immunity examination and wide spectrum vaccine development.

## Introduction

COVID-19 is bringing scenes of sci-fi movies into real life. Considering more than eleven million of already diagnosed cases and the growing number reported in the past few months, it seems to be far from over. The coronaviridae family also includes other respiratory syndrome causative agents in humans, the SARS-CoV, and MERS-CoV viruses^1^. The emergence of this viral pneumonia started in December/19 in the Hubei province, central China^2^, rapidly spreading around the globe, receiving the pandemic status at the beginning of March 2020. In the global actual scenario, there are almost 525.000 virus causative deaths, according to COVID-19 Dashboard by the Center for Systems Science and Engineering (CSSE) at Johns Hopkins University^3^ - *recovered at July 03, 2020*. Nevertheless, we should consider that there is not a regular correlation between the number of diagnosed individuals and mortality, among the affected countries, as shown in **Table 1**.

**Table 1.**
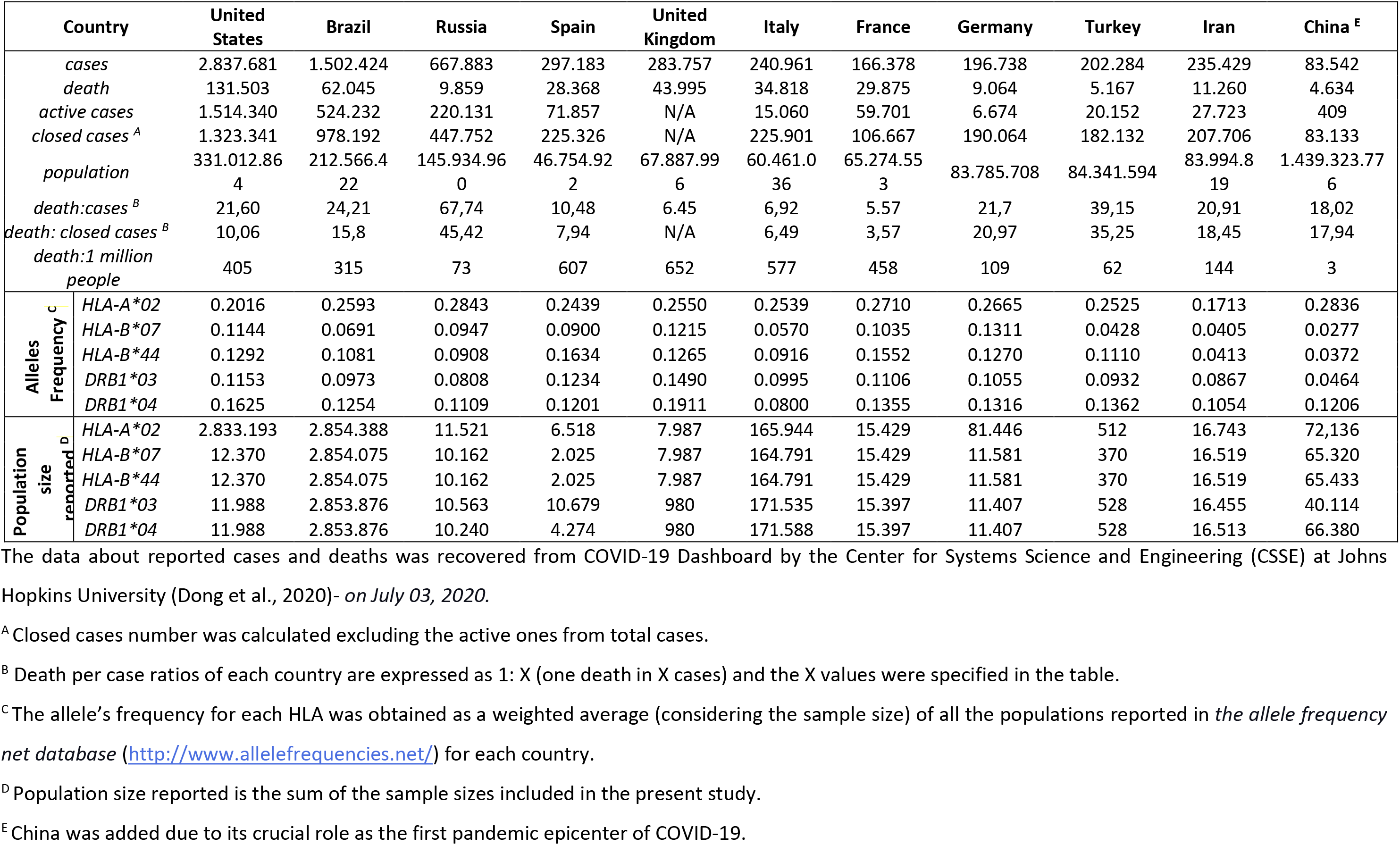
Alleles frequency information and data of the most affected countries considering the number of COVID-19 cases.

The United States, for example, shows the highest number of cases, which may be due to a better diagnosis rate, which influences the death rate calculation. In this context, Russia is the third country with a high number of cases, although its low mortality rate (one to 67.7 cases). Other countries with the most infected populations also present a low proportion of morbidity in diagnosed individuals as Germany (1:22). These values are quite different from those found in Italy (1:7) and France (1: 5.6), for example. It becomes even more alarming when we realize that each number is a human life.- *calculated data recovered at July 03, 2020*.

What elements could be dictating these differences? Social and cultural differences? The diverse starting point of government measures? Or genetic background presented by the different populations and SARS-CoV-2 strains around the world, causing distinct immune response profiles in critical patients? The present work will discuss this last question.

In the current pandemics, the first clues over the importance of cellular response are arising. For example, a work from Diao et al. (2020) found a negative correlation between CD4+ and CD8+ cell counts and disease severity in 522 patients from Wuhan, with laboratory-confirmed COVID-19. This fact places a pool of especial proteins orchestrating cellular immune mechanisms, as central players. Most of their genes are located on MHC locus. The MHC is the most polymorphic locus in the human genome^4^. This great variability enables that animal species could be able to fight, especially in a populational level, with the comparable mutational potential of pathogens, as viruses. Briefly, it is involved with the processing and presentation of small peptides in cell surfaces, derived from intracellular proteins, allowing the immune system to discriminate self from non-self, thus eliminating pathogens and tumors. At this point, a critical question is raised. We mentioned the MHC locus intentionally, not only MHC genes, considering that other proteins, fundamental to produce true epitopes, belong to this genomic region. Many studies, aiming to prospect tumoral or vaccine targets, focuses its prediction only on the pathogen or cancer sample MHC ligandome^5^. A potential to bind to different alleles do not confer to peptides their full potential to be a T cell epitope. A full T cell synapsis triggering demands additional requirements such as epitope immunodominance and pMHC:TCR physicochemical complementarity. Thus, *in silico* analysis considering additional steps on the antigen processing pathway and comparisons among putative targets and immunogenic epitopes, could present a better performance to prospect actual T cell epitopes, as in the current situation where no previous information is available.

Presenting a central role in this process, the HLAs (peptides presenting molecules in humans) constitute a link between the immune system surveillance and intracellular space status. It is known that different HLA alleles can bind and present a specific viral protein region (peptides) with altered efficiencies, which could provoke both susceptibility or resistance to a disease caused by a specific pathogen, depending on the type of MHC that the individual possesses.^6,7^

Thus, some HLA allotypes are unable to present some immunodominant epitopes (those responsible to initiate T-cell responses) from a specific virus, precluding the detection and fighting by the immune system. The opposite also occurs, the existence of HLAs with improved potential to present optimal viral targets, allowing the infection control^8^. The HLA alleles frequency vary among different populations. Some researches suggest that HLA alleles conferring pathogen resistance are most prevalent in areas with endemic diseases.

An important point to highlight in the current situation is that, even if the allele frequencies do not directly correlate with the percentage of the infected individuals on affected countries, what it can be explained by the probably enhanced affinity of SARS-CoV-2 spike protein by the ACE2 receptor^9,10^, the same cannot be said about differential outcomes of critical patients, without a deep HLA genotyping and investigation.

One can argue that this has already be done, in the 2003 SARS outbreak, with few of none significant associations. Nevertheless, we should consider that the number of genotyped individuals in that situation was extremely low compared with the current pandemic. Beyond that, many volunteers in those studies were health workers with putative contact with the virus^11^. Now, unfortunately, we are facing a scenario where we could extract more reliable correlations with alleles indicating better/worse prognosis in COVID19 patients. Large populational samples are essential in studies where we are dealing with a vast number of alleles (variables), as indicated above, contributing to minor effects, which could be important at a global level.

## Results and Discussion

### The HLA frequency relation and its apparent effect in COVID-19 immune responses

The virus’s ability to infect and spread in its pandemic should be related to a shared common global characteristic, as mentioned before. So, it is clear that the infection propagates disconnected from populations’ habits and origins, but the lethality varies between them. Important information arises from attending for the most frequent/rare MHC alleles in the most affected regions. The frequencies of these different HLA alleles vary widely among the countries with the highest number of cases ^12^. It may indicate a variation in the infection responses, which can influence morbidity. A close examination pointed out that none of the alleles frequency seems to be able to explain the death rate disparities independently.

Aiming to verify the alleles frequencies influences in disorder outcomes, and in the pandemic picture, we incorporated them into a Hierarchical Clustering Analysis (HCA) using the pvclust package at the RStudio platform. The inclusion of specific human leukocyte antigens (HLA) was refined based on previous studies references and its frequencies on the alleles data bank (considerable prevalence) distributed among some of the more predominant countries in terms of COVID-19 cases, plus the country where occurs the first epicenter of pandemics. Starting from 40 alleles we were excluding them by considering an absence of frequency information for some of the investigated countries and the preliminary HCA results. The final alleles selection includes four allotypes: HLA-B*07, HLA-B*44, HLA-DRB1*03, and HLA-DRB1*04. Each of them is a representative member of a distinct supertype (determining differential recognition and responses in the immune system). The HLA alleles DRB1*03 and DRB1*04 were already described as involved with autoimmune reactions elicitation ^4^. In 2006, some works detected autoantibodies directed against lung epithelial cells antigens, which could be mediating tissue damage, in some stages of the disease^13^. In this sense, alleles from the DRB1 gene may play a role in presenting targets from the virus sharing features with self-antigens. Furthermore, some studies highlighted the role of B*07 and DRB1 alleles in the development of SARS susceptibility and resistance^14^, and described HLA-B*44 as a predictive element in Hepatitis C Virus clearance^15^. Even considering the prevalence and importance of HLA-A*02 supertype, it was not included in HCA analysis considering its low contrast ratio between the countries. The values used in the HCA were the weighted average of serotype frequencies from all populations belonging to each considered country. **Table 1** summarizes the cited data. Additional negative symbols were artificially attributed to DRB1*03 values intending to detach its alleles frequencies in the HCA. This action was a result of a previous visual inspection correlating its subtle prominence in countries presenting a worse prognosis, classifying the allele as a potential determinant factor.

**Figure 1** shows the result for the HCA. The formed clusters seem to explain (in a general way) the global pandemic scenario with exciting results that indicate active contribution of the selected alleles and its consequent impact on how the COVID-19 affects countries population. Italy and France are the two more closely related countries, forming a ternary group with Spain. They have, respectively, a mortality index for closed cases of 1 to 6.49, and 1 to 3.57, while in Spain it is from 1:7.94. Taking into account the active cases the death per case rate decreases, being of 1:6.92, 1:5.57, and 1:10.48, in Italy, France, and Spain, respectively (*all values were calculated considering numbers recovered by July, 03*) 3 This group comprises three relevant nations considered as former significant pandemic epicentres, presenting high lethality indexes. A related cluster includes the United States, the United Kingdom, and Brazil. Currently, these countries represent a prominent alarming status, possessing an elevated contagious level. Its estimations of death per closed cases and death per general cases show quite a difference (from 1:15.76 to 1:25.21 in Brazil; 1:10.06 to 1:21.58 in the US), indicating they are in their epidemic course. The UK has no available information about the closed cases, only the death per case index (1 to 6.45), contrasting with the other nations in the cluster. Considering the deaths per million inhabitants, Brazil, the US, and the UK show values of 306, 401, and 652, respectively. These values are similar to the related cluster containing Spain (607 death/1M population), Italy (577), and France (458) which also have elevated numbers, indicating the high level of virus spread in all these countries. It is even more evident if we compare them to China, where the pandemic remained more concentrated in a specific location (Wuhan), and the country shows an index of three deaths per million people.

**Figure 1.**
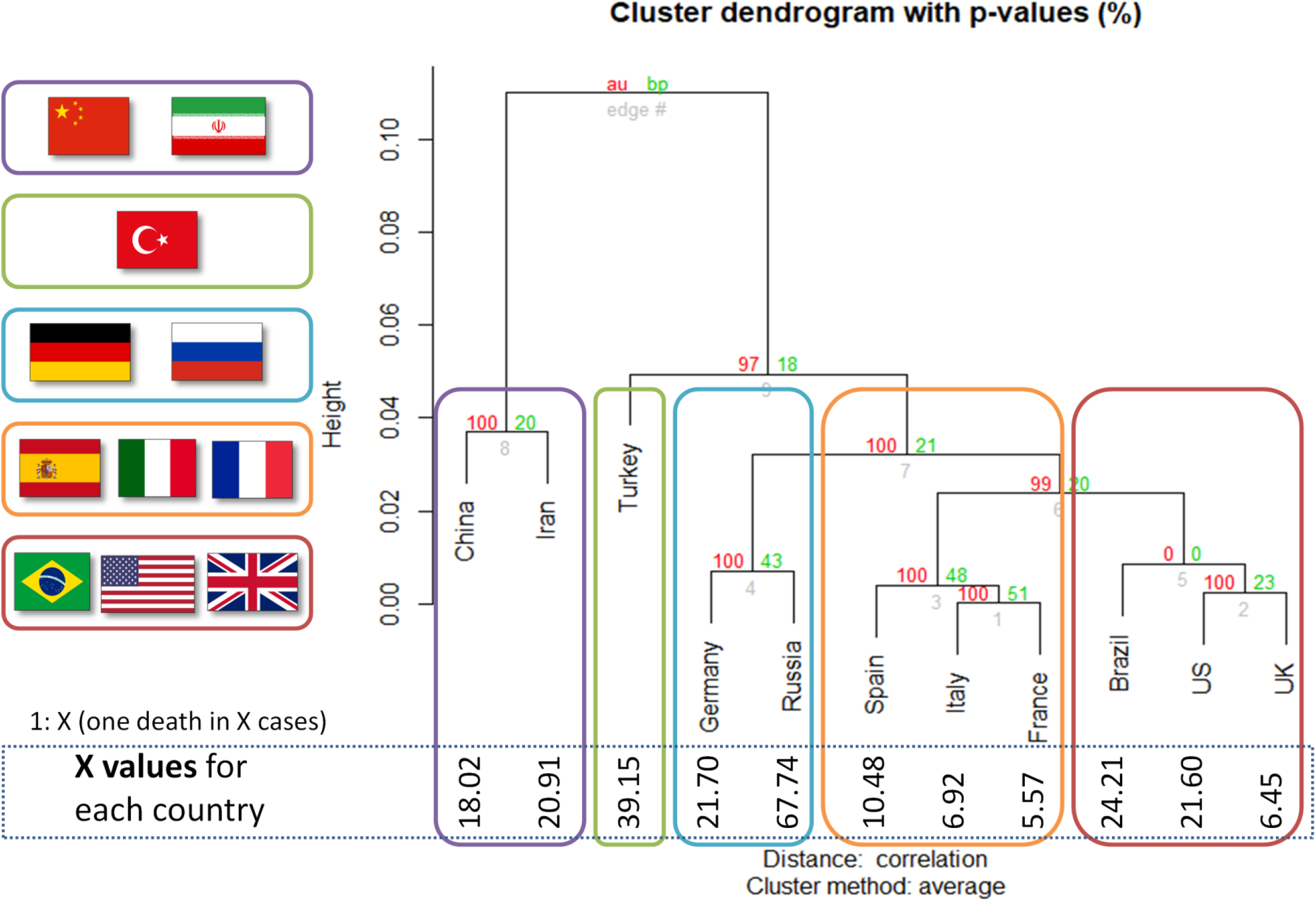
Hierarchical Clustering Analysis from the most affected countries in number of COVID-19 cases reported, based on its HLA frequencies. The image represents the HCA result for a clusterization concerning the frequencies of the following alleles: HLA-B*07, HLA-B*44, HLA-DRB1*03, and HLA-DRB1*04. Four clusters comprising countries with similar death rates are showed. High-rates countries as Spain, Italy, and France grouped. In the same way, low-rate nations like Germany and Russia fall in the same branch. Its required frequencies were obtained from Alleles Frequency Net Database (http://www.allelefrequencies.net/) and are also compiled in Table 1. Values at branches are AU p-values (left), BP values (right), and cluster labels (bottom). Clusters with AU ≥ 95 are indicated by the rectangles. Death per case ratios of each country are expressed as 1: X (one death in X cases). The calculated values for “X” are depicted in HCA.

The next cluster includes Germany and Russia. These two countries present more bland pandemic indexes. The death per closed cases in Germany is similar to death per general cases (1:20.97 and 1:21.7, respectively), it does not occur in Russia (1:45.42 and 1:67.74) due to a large number of active cases. Both nations have a more favorable score of death per million people if compared to the cited above countries (108 in Germany and 71 in Russia). Turkey branches apart from the above-indicated nations, with favourable values of the considered indexes (1:35.25 and 1:39.15). China (1:17.94 and 1:18.03) and Iran (1:18.45 and 1:20.91) fell in a separate group, having very similar death rates, despite their ethnic divergences (**Table 1**).

Trying to establish an initial validation of the noticed alleles effects, we include Pakistan, Portugal, and Peru (low-rate mortality indexes), and Romania, in four independent clustering approaches (**Table S1 and Figure S1**). Portugal (**Figure S1A**), Peru (**Figure S1B**), and Pakistan (**Figure S1C)** cluster with other nations (Turkey, China, and Germany, respectively) presenting low-rate death indexes. Romania grouped with Brazil (**Figure S1D)**, both countries have death per closed case relatively high. As in the clustering presented in Figure 1, the groups depicted here share aspects of disease severity, indicating that the selected alleles (HLA-B*07, HLA-B*44, HLA-DRB1*03, and HLA-DRB1*04) are good predictors. However, it does not prevent the inclusion of additional alleles or elements to improve the predictability, on the contrary.

### HLA-A*02:01 as an example of universal protective alleles in human populations

An alternative hypothesis is that the T cell response governing SARS-CoV-2 recovering and clearance is through prevalent alleles in populations, as the HLA-A*02 supertype, which could be a good sign. Initially, it seems to be a contradictory reasoning, but in light of the mortality rates in other SARS causative viruses (around 10% or above)^16^, the current values may indicate a more effective worldwide cellular response. This idea is supported by many studies reporting HLA-A*02:01 as a pivotal allele in viral infections. A consultation on Immune Epitope Database (www.iedb.org) returned 849 references describing positive T cell response against viral peptides presented in the context of HLA-A*02:01 allele (accessed on May 26, 2020). Besides, 20 out of 34 immunogenic epitopes described for coronaviruses are restricted to HLA-A*02:01. In 2003 pandemics, lymphocytes from previously infected individuals were able to eliminate cells presenting SARS-CoV epitopes restricted to HLA-A*02:01 molecules^11,17^ up to six years later from recovering^18^, demonstrating the importance of this allele on viral clearance and T cell central memory. A conducted study with individuals from China and Hong Kong evidenced that more than half of the SARS recovered subjects were HLA-A*02:01 positive^19^. It is noteworthy, considering that the frequency of this allele is 0.1090 and 0.0620 in China and Hong Kong, respectively. The HLA-A*02:01 frequencies among the top 10 infected populations is high (median 23.27).

In our analysis, most of the recovered HLA-A*02:01 epitopes described in past epidemics seem to present conserved sequences compared to their equivalent in SARS-CoV-2 (**Table 2**), as described in other works ^20,21^. Nevertheless, in case of discordant peptide sequences, the simple sequences comparison of SARS-CoV and SARS-CoV-2 may provide us little information about the impact of these alterations on the immunogenic potential on current putative viral targets. The analysis of structural and physicochemical features in the peptide-MHC (pMHC) surfaces that contact the T cell receptors, considers the combined molecular elements involved in T cell activation. Such structural investigation has already demonstrated its potential, explaining differential immunogenicity among epitopes from diverse viral strains or tumoral origins ^22,23^. Since usually there is no available crystal for all specific targets complexed in HLA-A*02:01 allele, we construct all customized complexes through our reliable DockTope tool for pMHC modeling (http://tools.iedb.org/docktope/) ^24^. This structural analysis gives us two alternative scenarios: the TCR interacting surfaces of both pHLA-A*02:01 complexes were similar **(Figure 2A)**, pointing out for preserved immunogenicity in SARS-CoV-2 targets; or the targets presented subtle physicochemical alterations in complexes harboring SARS-CoV-2 peptides, compared to former SARS-CoV. Amazingly, in this case, some of them turned into closely related surfaces presented by previously immunogenic epitopes, from non-related viruses **(Figure 2B)**. In the depicted example, the SARS-CoV-2 peptide SIIAYTMSL demonstrates structural convergence of physicochemical features with the immunodominant epitope M1_58-56_ GILGFVFTL, from the Influenza virus. Both epitopes share 2/9 amino acids, reinforcing the importance of structural investigation to prospect cross-reactive targets. In our analysis, to make it evident that the comparisons were not result from structural biases, we provide a small sample of unrelated models from our CrossTope database (www.crosstope.com) ^25^, containing TCR interacting surfaces from HLA-A*02:01 structures presenting immunogenic epitopes **(Figure S2**). These first comparisons uncovered interesting scenarios. Firstly, even those peptides with sequence alterations in SARS-CoV-2, but without molecular modifications in the TCR interacting surfaces, remain good candidates to immunization strategy. The examples where SARS-CoV-2 peptides shown subtle alterations compared to their corresponding SARS-CoV targets could indicate a change in immunogenicity or even a complete loss of it. However, in this situation it resembles highly immunogenic epitopes from other viral organisms. This evidence emphasizes the need to investigate this new face of the immunogenic prism.

**Table 2.**
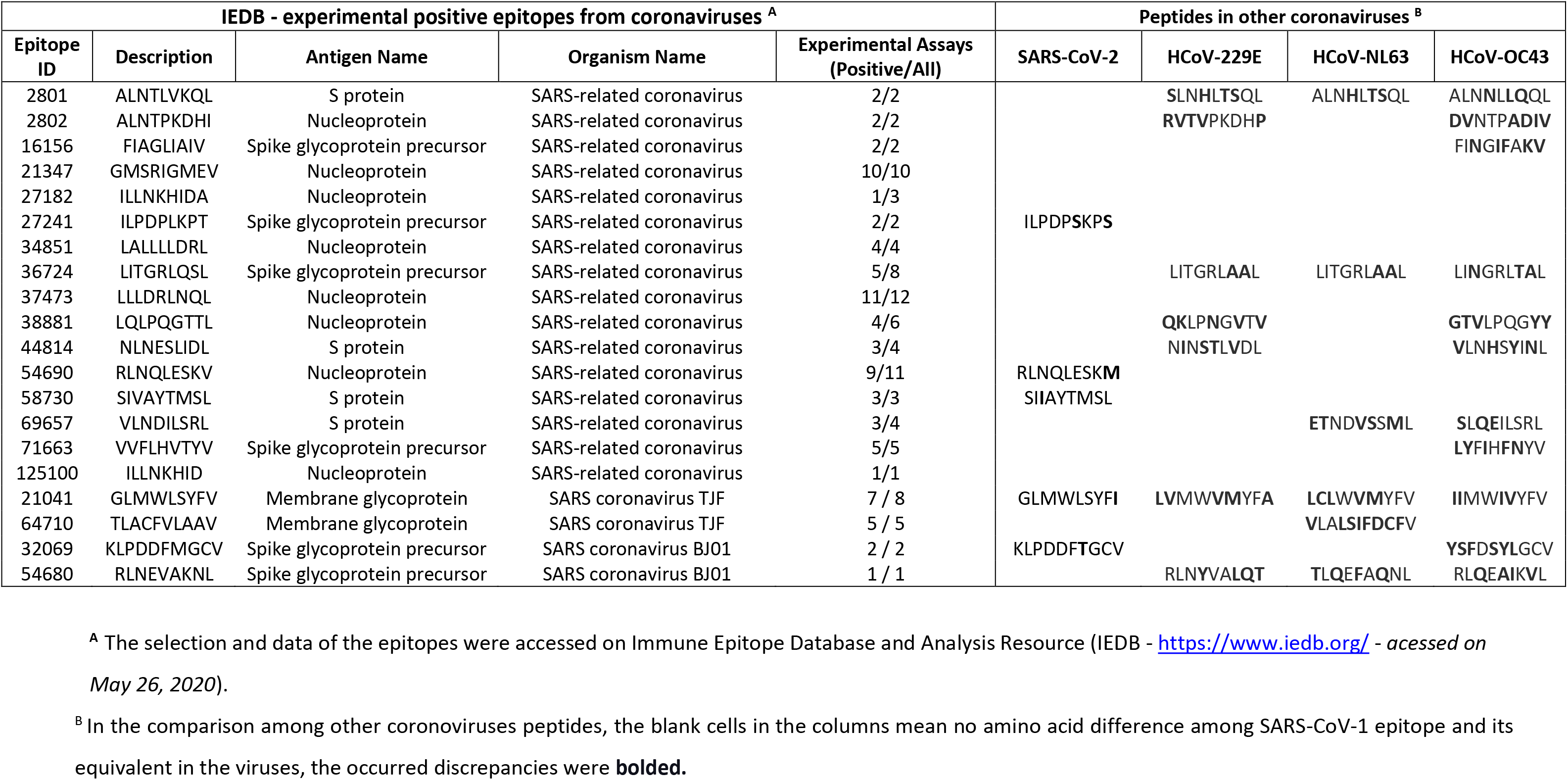
Recovered HLA-A*02:01 epitopes from SARS-CoV, SARS-Cov-2 and other alphacoronaviruses.

**Figure 2.**
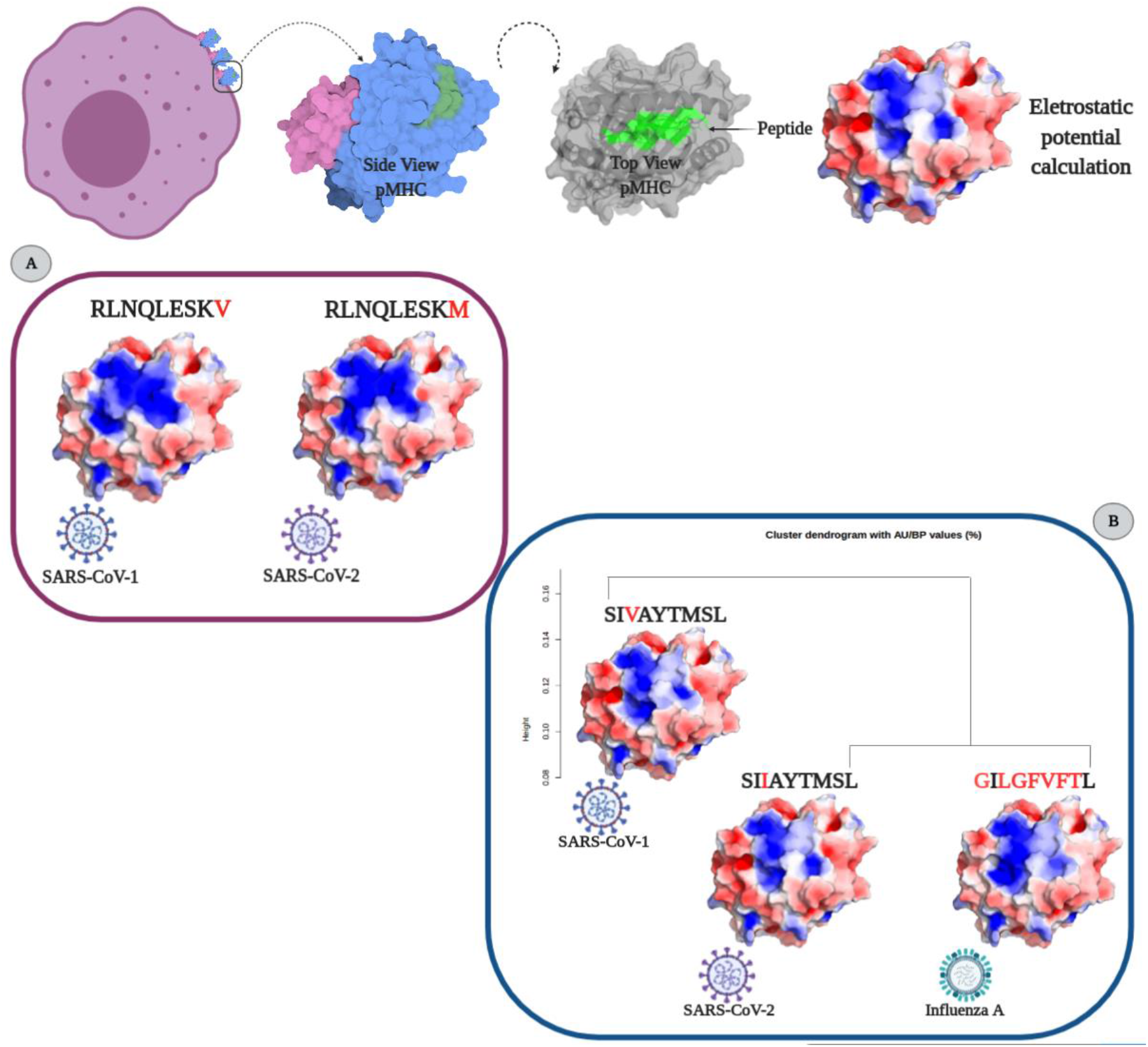
Structural analysis of SARS-CoV HLA-A*02:01 epitopes. A sequence demonstrating the TCR-interacting surface is depicted above. From left to right we can observe 1) the MHCs in the cell membrane; 2) a side view of pMHC with alpha chain in pale blue, B2-microglobulin in pale purple and the peptide in green; 3) a top view of pMHC with the peptide region highlighted in green and 4) the pMHC surface with electrostatic potential distribution and topography, the main elements involved in immune response stimulation. Negative values are represented in red and positive ones in blue. A) SARS-CoV peptide sequences showing high similarity in sequence and structural features. B) SARS-CoV SI**V**AYTMSL/SI**I**AYTMSL sequences presenting subtle differences. An HCA analysis with the IAV **G**I**LGFVFT**L epitope demonstrates that it is even more similar to SIIAYTMSL SARS-CoV-2 peptide sequence.

### Searching for SARS-CoV-2 shared immunogenic fingerprints in targets from HCoVs and other prevalent viruses in human populations

The observed molecular similarity between pMHCs complexes containing peptides from SARS-CoV-2 and Influenza viruses brings us toward another attractive hypothesis that refers to a universal previous cytotoxic response present in populations from all over the world, triggered by previous infections. The first suspects to investigate were past contact with targets from remaining betacorononavirus (OC43) and alphacoronavirus genus members (229E and NL63). Epidemiological studies reported that around 15-30% of the common cold are caused by this group of pathogens^26^. Even considering that they are viruses with a zoonotic origin, we would expect many spillover events throughout the history of humans, maintaining regular contact with our species^27^. A codon usage analysis, involving BCoV and HCoV-OC43, suggests that an ancestor coronavirus could be present even 200 kyr ago, in early men^28^. Therefore, we would expect that this group of pathogens has also contributed to shaping our current immune system repertoire. Guided by this supposition, we compared the immunogenic SARS-CoV epitopes with 229E, OC43, and NL63 corresponding protein sequences, looking for shared elements involved in immunogenicity triggering. Such analysis presented a clear example where sequence comparison might be hiding shared patterns not detectable by single amino acid identity alignment. In **Table 2**, the sequences identities ranged around 50%, a value usually not detected by regular alignments methods prospection. Nevertheless, when we inspect these same epitopes in the context of pMHC structural models harboring these peptides sequences from SARS-CoV-1, SARS-CoV2, and alphacoronavirus in HLA-A*02:01 alleles, intriguing fingerprints arose. A similar electrostatic distribution and topography, on the TCR interacting surfaces from the pMHCs, can be observed among SARS-1, SARS-2, and other coronaviruses members (229E and OC43) (**Figure 3A**). It is important to reinforce that both 229E and OC43 putative epitopes were predicted as strong binders to HLA-A*02:01 (data not shown), strengthening their potential as actual triggers for SARS-CoV cross-reactivity. The peptides from the beta-CoVs are quite similar (**GLMWLSYF**L and **GLMWLSYF**V, between SARS-1 and 2, respectively). However, the peptides from 229E (LV**MW**VM**YF**A) and OC43 (II**MW**IV**YFV**) are distinct, evidencing the importance of the structural investigation. Furthermore, other peptides derived from alpha-CoV viruses presented a less prominent similarity in physicochemical features to immunogenic SARS-CoV-1 epitopes (data not shown). However, they are also potential targets to investigate. A work recently deposited in bioRxiv showed that 34% SARS-CoV-2 of seronegative healthy individuals presented S-reactive CD4 T cells. These cells react almost exclusively with the C-term epitopes region, characterized by higher homology with spike protein of human endemic common cold coronaviruses. Nevertheless, none of the putative cross-reactive epitopes are pointed-out, nor the structural basis hypothesized, but it reinforces our propositions^29^. Evidences of many CD4+/CD8+ cross responses against many SARS-CoV proteins in unexposed individuals were extensively described in Griffoni et al. ^20^, without the specific identification of sequence targets. Other work describing correlations of CD4+/CD8+ T cell differential phenotypes between acute (highly activated cytotoxic) and convalescent (stem-like memory) patients, was conducted by Karolinska COVID-19 Study Group^30^. Interestingly, they described the occurrence of SARS-CoV-2-specific T cell responses elicited in the absence of circulating antibodies in non-infected individuals, suggesting that previous contacts could be the triggers of these cross-reactive events.

**Figure 3.**
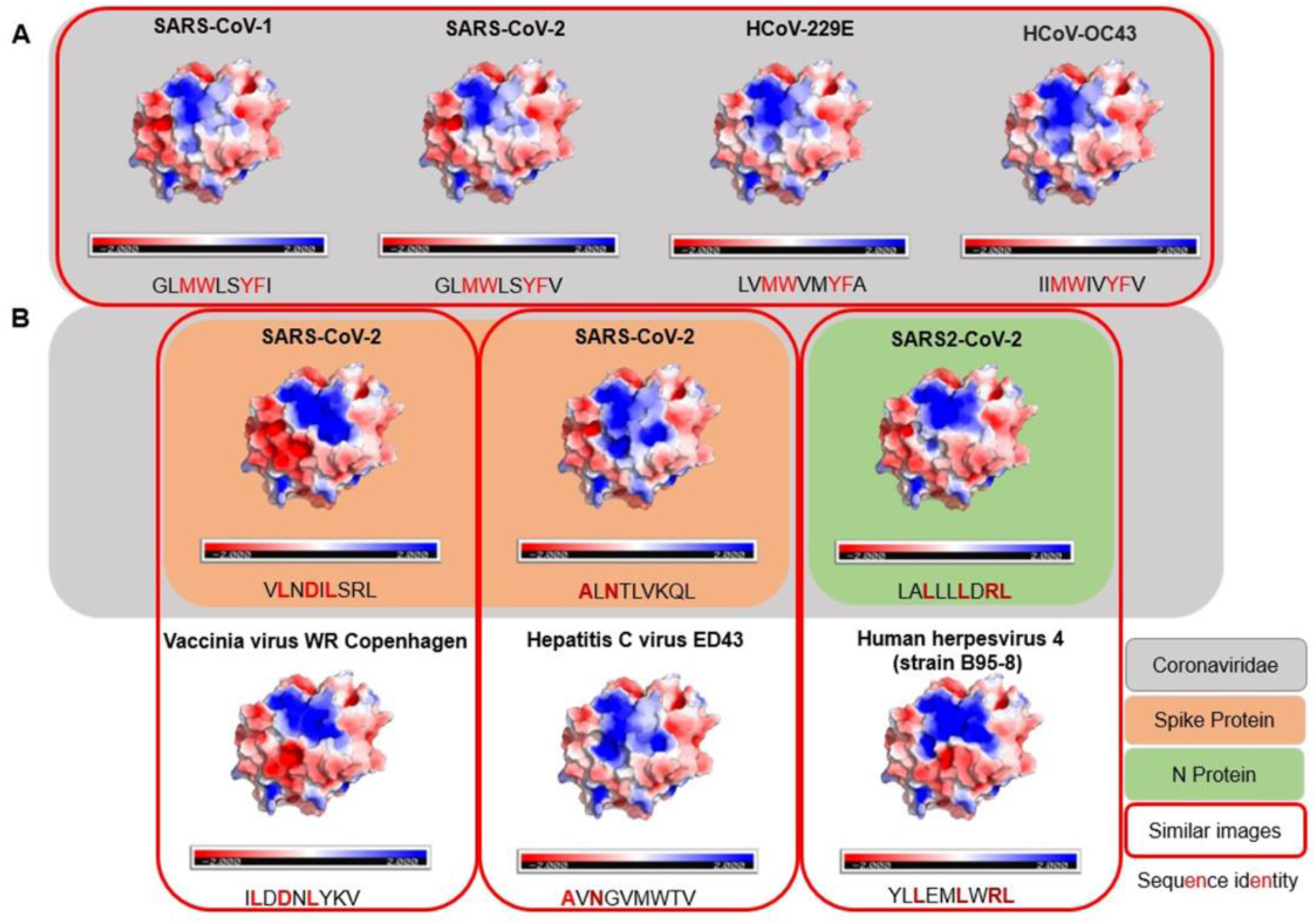
Comparison of electrostatic surface distribution and topography of SARS-CoV peptides with alphacoronaviruses and other prevalent viruses in human populations. pMHCs models were compared in terms of topography and electrostatic distribution. In A, two beta (SARS-CoV) and two alpha (229E and OC43) coronaviruses show similar electrostatic distribution and topography despite its sequence divergences. In B, a panel of three SARS-CoV peptides is compared to pMHCs containing viral epitopes from members of other families. An unexpected shared structural similarity arises, disregarding their lack of sequence identity and phylogenetic relationship. Electrostatic calculations are represented as negative (red) and positive (blue) charges. Sequence identity is depicted as red letters in the peptide sequences.

In this regard, given the previous identification of a similar target from the Influenza virus (M1_58-66_ GILGFVFTL) with a SARS-CoV epitope, the next step was to scrutinize other trigger candidates on previous described viral epitopes. To perform the comparison, we recover pMHC structures on CrossTope T cell epitope databases (*http://crosstope.com/*^25^) looking for immunogenic fingerprints common to SARS-CoV-1 epitopes and unrelated viruses. As demonstrated before, considering that SARS-CoV-1 and SARS-CoV-2 peptides are sequence and structurally related, the comparison with experimentally described epitopes from SARS-CoV-1 seems to be more appropriate. The results from these comparisons were extraordinary. When we look for pHLA-A*02:01 structures, not only the previous example of M158-66 IAV epitope matched with SARS epitopes, but 13 out 20 CD8+ coronavirus epitopes, recovered from Immune Epitope Database, has counterparts in targets from common circulating viruses, concerning depicted molecular features. **Figure 3B** presents three examples of SARS-CoV peptides presenting stunning structural identity with viral epitopes. The matched targets belong to viruses from three different families (Herpesviridae, Poxviridae, and Flaviviridae). It is important to think that these targets would probably not be investigated and selected in an approach using regular methods, given that no apparent identity is evidenced by any of these structurally related epitopes with SARS-CoV sequences. The remaining comparisons can be viewed in **Figure S3**. Importantly, when we consider these images correspondences, the comparisons with pMHCs from unrelated viruses were more conspicuous than those from other representatives of HCoVs. It seems a paradox, given the natural expectation (considering its phylogenetics proximity) of a more intimate relation between alpha and beta CoVs peptides with SARS-CoV epitopes.

## Concluding Remarks

In COVID-19, the aetiological agent, SARS-CoV-2, is well established. Nevertheless, the same cannot be regarding all elements participating in severe acute respiratory syndrome (SARS) etiology. For this purpose, two aspects of cellular response were here approached: the impact of the MHC allelic constitution of populations on COVID-19 outcome and the structural analysis of immunogenic elements presented by SARS-CoV and putative SARS-CoV-2 T cell epitopes.

In the first section, an HLA allele frequency investigation was conducted on some of the most affected nations (considering COVID-19 cases number) to elucidate populations’ genetic elements probably involved in lethality indexes. By only using four frequencies of prevalent alleles, we were able to cluster countries presenting similar death rates profiles concerning both closed and active cases. It seems to contribute with the determination of population response facing COVID-19, in addition to the political and medical interventions which are, in every country context, also extremely essential. It is necessary to emphasize that this approach prototype was just a primary prospection of how these components could operate in complex scenarios. May have countless more factors (and alleles, probably in haplotypes) involved with the mortality determination ratio. Nonetheless, it highlights the importance of a worldwide genotyping effort to clarify this landscape.

The epitope structural analysis and their relationship with other HCoVs and unrelated viral targets unveiled noteworthy observations. These pieces of evidence may open two avenues of investigation. The high degree of molecular conservation between SARS-CoV-1 epitopes and its corresponding sequences in SARS-CoV-2 allows its use in vaccine development to stimulate cross-reactive responses, covering distinct SARS-CoV-2 strains, for example. In this regard, animal reservoirs should be inspected looking for beta CoVs with the potential to spill out of their natural hosts to humans. Peptides sequences from these putative HCoVs pathogens could be structurally compared searching for cross-reactive T cell targets to be used in a virtual future occurrence of a new coronavirus spillover phenomenon. Such preventive strategy could abbreviate steps to develop immunotherapeutic methods, avoiding the emergence of new pandemics. The second line of the investigation resulted in an even more attractive hypothesis. Previous infections with different alpha/beta-CoVs and unrelated common viruses can be generating memory T cells against SARS-CoV-2, in a significant portion of the population. This pool of cells in different individuals is providing a universal immunogenic shield against SARS-CoV-2 and, probably, against other potentially emergent and endemic viruses. This mechanism seems to be evolutionarily constructed by regular cross-reactive contacts. Moreover, the defense appears to be associated with prevalent alleles, which probably present peptides harboring fingerprints of immunogenicity shared by epitopes that regularly infect humans.

## Methods

### Alleles search and data evaluation

To start the investigation, we selected highly affected countries concerning the number of COVID-19 cases ^3^ (*recovered at July 03, 2020*), plus China, which was the first pandemic epicenter, and other selected countries to validate our analysis. The alleles having a frequency equal to or higher than 10% (in at least one of the selected countries) were examined. Some HLAs were rejected due to their lack of information in some selected countries or considering their similar frequencies across nations, which could give little information on the clusterization method. The data were recovered from The Allele Frequency Net Database (AFND)^12^, selecting the locus and allele of all populations for each specified country included in the analysis. A list of country populations, including sample size from where the allele frequencies were obtained, is provided. A weighted average was calculated for each allele to avoid size sample biases. A better description of the applied methodology can be visualized as a graphical scheme at **Figure S4**.

### Hiararchical Clustering

In the second step, the weighted average of B*07, B*44, DRB1*03, and DRB1*04 frequencies values for each country was used to fuel a hierarchical clustering analysis (HCA) on package Pvclust ^31^. The HCA allow the classification of several objects (countries) into some groups (clusters) according to similarities between them (allele’s values). Pvclust calculates probability values (p-values) for each cluster using bootstrap resampling techniques. Two types of p-values are available: approximately unbiased (AU) p-value and bootstrap probability (BP) value. Multiscale bootstrap resampling is used for the calculation of AU p-value, which has superiority in avoid bias over BP value calculated by the ordinary bootstrap resampling. Generally the AU p-value is a better estimator of the reliability of the clusters obtained. The bootstrap analysis was performed with the number of bootstrap replications being B = 10.000.

### SARS-CoV Epitopes propections

We prospected the IEDB database for experimental coronavirus epitopes, finding 20 immunogenic epitopes restricted to HLA-A*02:01 allele. Those epitopes were derived of the N (nucleocapsid) protein, Surface (spike) protein, and Membrane glycoprotein. To verify if those epitopes have a counterpart in the SARS-CoV-2 proteome, we used a Needleman-Wunsch Global Align Nucleotide Sequences (BLAST) using the protein sequences of the 2002/2003 SARS virus and the SARS-CoV-2 from Wuhan. To check if the binding affinity was preserved in discordant SARS-CoV-2 sequences, we predict it through NetMHCcons tool (http://www.cbs.dtu.dk/services/NetMHCcons/)^32^. The same tool was used for the remaining HCoVs peptides.

### HCoVs epitope prediction

We selected viruses from the Alphacoronavirus (229E and NL63), and Betacoronavirus (OC43), checking if these strains possess corresponding SARS-CoV epitopes that could generate similar surfaces, and consequently, act as triggers of cross-reactivity. To this, we screened the protein sequences of different HCoVs searching for some amino acid identity with immunogenic targets described for SARS-CoV-1. Then, they were modeled to verify if their surfaces of interaction with TCR present structural conservation throughout different coronaviruses.

#### Structural analysis

To generate customized peptides anchored in the desired MHCs (pMHC), all coronavirus epitopes were modeled in HLA-A*02:01 allele by Docktope Tool. The 3D models were used as input in the Pymol software (https://www.schrodinger.com/pymol) to calculate their electrostatic surfaces to verify how much the amino acid changes impacted the overall charge disposition in those models.

### Structural Hierarchical Clustering

We compared the pMHC complexes of both viruses with several other complexes from selected HCoV sequences and from those included in the Crosstope Database (www.crosstope.com), which harbors several previously described epitope sequences of immunogenic targets from the IEDB. All images of SARS-CoV and CrossTope HLA-A*02:01 epitopes were used to perform the comparisons. We utilized ImageJ^33^ to extract the Red-Blue-Green (RGB) values from electrostatic surface regions that contact T cell receptor. These regions were inferred by contact calculation of ternary crystals comprising p:HLA-A*02:01:TCR. The regions present colors that can be more positive, neutral or negative in that regards to the electric charges and this color information was converted in numeric values of mean, mode and standard deviation to feed the hierarchical clustering program pvclust, ran by the R Studio to perform this pairwise comparison and group the most similar surfaces together.

## Data Availability

The structures of pmhcs containing immunogenic epitopes can be found in the CrossTope database. The CoV targets were recovered on the immune epitope database.

http://crosstope.com/

https://www.iedb.org/

## Acknowledgments

We thank to National Council for Scientific and Technological Development (CNPq) and National Council for the Improvement of Higher Education (CAPES) for their support.

## Authors Contributions

E.C.A performed the structural analysis in cross-reactivity predictions. M.A.S.B recovered epitope and genome sequences of coronaviruses and performed sequence analysis. M.R.M conducted all the experiments regarding HLA frequencies and wrote the manuscript. G.F.V designed the study, coordinated the experiments and wrote the manuscript.

## Declaration of Interests

The authors declare no competing interests.

**Figure S1. *Additional Hierarchical Clustering Analysis based on countries HLA frequencies***. The figure depicts the inclusion of four countries: Portugal (A), Peru (B), Chile (C), and Romania (D) in separate HCAs. It illustrates the results for clusterization concerning the frequencies of the following alleles: HLA-B*07, HLA-B*44, HLA-DRB1*03, and HLA-DRB1*04. Values at branches are AU p-values (left), BP values (right), and cluster labels (bottom). Clusters with AU ≥ 95 are indicated by the rectangles. Death per case ratios of each country are expressed as 1: X (one death in X cases).

**Figure S2. *TCR interacting surfaces from pMHC immunogenic structures***. Nine pMHC from HLA-A*02:01 bound to experimental epitopes are represented. This panel of images was provided to evidence molecular signs diversity that can be found in these targets.

**Figure S3. *Remaining comparison of SARS-CoV epitopes and immunogenic epitopes from CrossTope Database***. Shared fingerprints are also observed in the above-exemplified pairs of structures indicating a general structural signature.

**Figure S4. *Resume the method applied in “The HLA frequency relation and its apparent effect in COVID-19 immune responses” section***. The weighted average (WA^1^) of each allele’s frequency was obtained for the selected countries at *the Allele frequency net database* (*AFND*). It was used in a hierarchical clustering analysis (HCA) through the pvclust package in the R-Studio software.

## Notes

### Competing Interest Statement

The authors have declared no competing interest.

### Funding Statement

This work was supported by the National Council for Scientific and Technological Development (CNPq) and National Council for the Improvement of Higher Education (CAPES) for their support.

